# Statistical and network-based analysis of Italian COVID-19 data: communities detection and temporal evolution

**DOI:** 10.1101/2020.04.17.20068916

**Authors:** Marianna Milano, Mario Cannataro

**Author notes:** To whom correspondence should be addressed:{, }.

## Abstract

Coronavirus disease (COVID-19) outbreak started at Wuhan, China, and it has rapidly spread across China and many other countries. Italy is one of the European countries most affected by the COVID-19 disease, and it has registered high COVID-19 death rates and the death toll. In this article, we analyzed different Italian COVID-19 data at the regional level for the period February 24 to March 29, 2020. The analysis pipeline includes the following steps. After individuating groups of similar or dissimilar regions with respect to the ten types of available COVID-19 data using statistical test, we built several similarity matrices (reported in Supplementary file). Then, we mapped those similarity matrices into networks where nodes represent Italian regions and edges represent similarity relationships (edge length is inversely proportional to similarity). Then, network-based analysis was performed mainly discovering communities of regions that show similar behaviour. Then, network-based analysis was performed by running several community detection algorithms on those networks and by underlying communities of regions that show similar behaviour. The network-based analysis of Italian COVID-19 data is able to elegantly show how regions form communities, i.e. how they join and leave them, along time and how community consistency changes along time and with respect to the different available data.

## 1 Introduction

Coronavirus disease, called COVID-19, emerged in the city of Wuhan, in China, in November 2019 [9].

The disease is caused by the novel coronavirus Sars-CoV-2 [10] and its clinical manifestations include fever, cough, fatigue, chest distress, diarrhoea, nausea, vomiting [2] and also acute respiratory distress syndrome in severe cases [8].

COVID-19 is featured by a long incubation period, high infectivity, and different transmission ways [4]. The contagion happens mainly through respiratory and blood contact with the coronavirus.

In a few months, COVID-19 epidemic quickly spread to Asian countries and it reached more than 200 countries in the world, causing about tens of thousands of deaths.

On March 11, 2020, COVID-19 disease was declared a pandemic by the World Health Organization.

In Italy, COVID-19 has been identified on January 2020 [5] and the outbreak started in Lombardia and Veneto at the end of February 2020. From the northern regions of Italy, the disease spread very quickly to the nearest regions and then to the rest ones. Italy has considered one of the main epicentres of the pandemic recording a number of infected equal to 97689 and a number of deaths equal to 10799 up to 29rd of March. In this paper we want to underline difference at regional level and to evidence how the disease has spread along time. To do this, both statistical and network-based analysis on Italian COVID-19 data has been performed. The data are daily provided by the Italian Civil Protection, and they refer to the period February 24 to March 29, 2020. In particular, we analyzed the Italian COVID-19 data at the regional level in order to evaluate which regions show a similar trend. Then, for each type of data, we built the related networks where the nodes represent the Italian region, and the edges represent a level of similarity among them. Finally, we evaluate how the networks evolve over the weeks up to the end of March 2020. The main contribution of the the paper is a network-based representation of COVID-19 diffusion similarity among regions and graphbased visualization to underline similar diffusion regions. The rest of the paper is organized as follows: Section 2 discusses the Italian COVID-19 data, the statistical and network-based analysis, Section 3 discusses the results, Finally, Section 4 concludes the paper.

## 2 Material and Methods

In this section, we investigate the evolution of the coronavirus pandemic in Italy from a statistical and network-based perspective, The analysis uses data at different temporal zoom e.g. by analysing the period from February 24 to March 29, 2020 and by focusing on single week as well as the entire observation. Moreover analysis comparing data at regional level is also provided. The analysis pipeline includes the following steps:

1. Building of ten similarity matrices (one matrix for each data measure. The (i,j) value of the matrix for data k (e.g. Swab data) represents the *p-value* of the Wilcoxon statistical test obtained by performing the test on the swab measures of region i with respect to region j. Lower *p-value* means that regions are more dissimilar with respect to that measure. Higher *p-value* means that regions are more similar with respect to that measure. We used the usual significance threshold of 0.05, thus matrices report only *p* − *vales >*= 0.05, while *p* − *values <* 0.05 are mapped to zero. In such a way the *p-value* is used as a similarity measure.
2. Mapping similarity matrices to networks. We map each matrix M(i,j) to a network N, where nodes represent the Italian regions and an edge connects two regions (i,j) if the *p-value* in the similarity matrix is greater than the threshold. Edges are weighted with the *p-value*. This allows to draw edges with length inversely proportional to similarity.
3. Temporal analysis of networks. For each data measure, the corresponding graph at different time points (i.e. at end of week 1,2, …, 5) and for all the time period are compared. In current analysis, the network at week k, is built considering only the time points collected in that week k and not considering past weeks. In future work we plan to consider aggregated data, i.e. data at week k, includes all data from week 1 to k.
4. Community detection. For each network we extracted subgroups of regions that form a community on the basis of similarity point of view. The identification of community is performed on the networks related to the observation period and for all single week. The, we analized the evolution of the communities at different time points, i.e. at the end of the first week, after tree weeks and after five weeks (the observation period).

### 2.1 Input DataSet

The present analysis was carried on the dataset on COVID1-9 updated in https://github.com/pcm-dpc/COVID-19 database, provided by the Italian Civil Protection. The dataset consist of:

- Hospitalised with Symptoms, the numbers of hospitalised patients that present COVID-19 symptoms;
- Intensive Care, the numbers of hospitalised patients in Intensive Care Units;
- Total Hospitalised, the total numbers of hospitalised patients;
- Home Isolation, the numbers of subjects that are in isolation at home;
- Total Currently Positive, the numbers of subjects that are coronavirus positive;
- New Currently Positive, the numbers of subjects that are daily coronavirus positive;
- Discharged/Healed the numbers of subjects that are healed from the disease;
- Deceased, the numbers of dead patients;
- Total Cases, the numbers of subjects affected by COVID-19;
- Swabs, the numbers of test swab carried on positive subjects and on subjects with suspected positivity.

The data are daily provided for each Italian region. The data occupies 47.6 Mbytes of memory.

### 2.2 Statistical analysis

We analyzed the trend of each type of data for the period February 24 to March 29, 2020. We computed the main descriptive statistics for all regions in the study period. All analysis are performed by using R software [6]. The Figure 1 conveys the evolution of all dataset over days.

**Figure 1:**
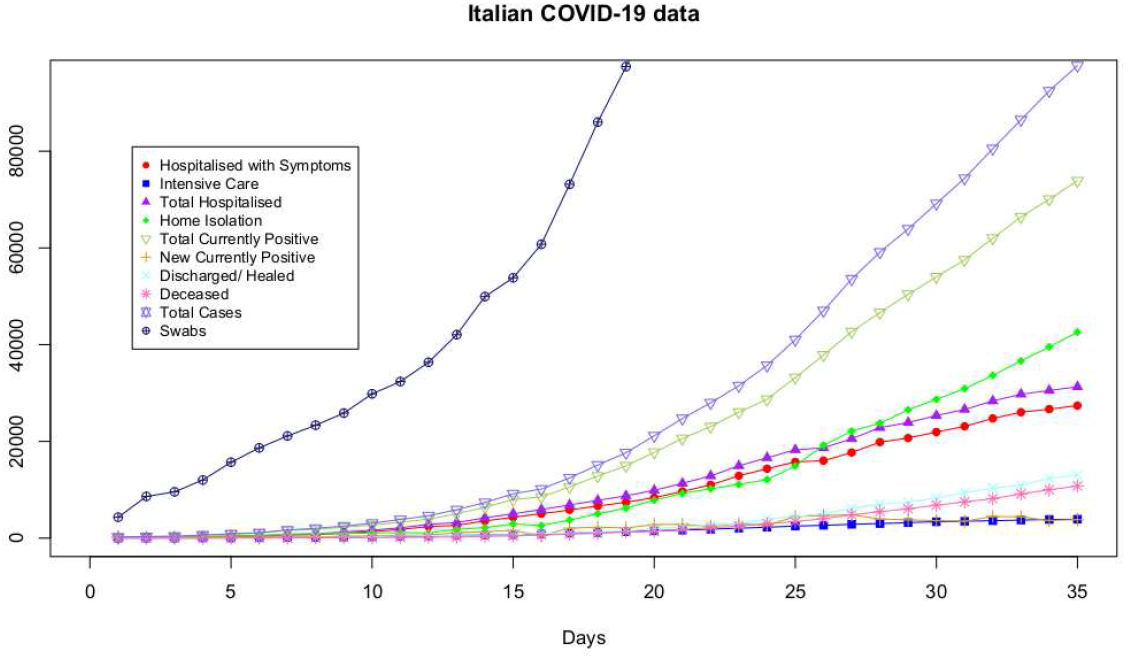
The figure show the trends of Hospitalised with Symptoms data, Intensive Care data, Total Hospitalised data, Home Isolation data, Total Currently Positive data, New Currently Positive data, Discharged/ Healed data, Deceased data, Total Cases data, Swabs data. Day 1 is February 24, 2020.

Then, we analyze the data trends by considering each single week. The first week starts on February 24 and ends on March 1, the second week starts on March 1 and ends on March 8, the third starts on March 9 and ends on March 15, the fourth starts on March 16 and ends on March 22, the fifth starts on March 23 and ends on March 29.

As a preliminary test, we applied Pearson’s chi-square test. Since *p-value* was less than 0.05 for each distribution, we decided to use non-parametric test for the following comparison. As initial step, we used the Wilcoxon Sum Rank test to carry out an analysis within the same type of data for all weeks and then, for each single week. The Wilcoxon test is a non parametric test designed to evaluate the difference between two treatments or conditions where the samples are correlated. The Wilcoxon test performs a pair-wise comparison among regions with the goal to evidence which ones show different trend. For this reason, we built a similarity matrix for each couple of regions, for each of the available COVID-19 data. Table 1 reports the similarity matrix related to Hospitalised with Symptoms network in the observation period. We reported the all similarity matrices computed for Italian COVID-19 data in the observation period e in the sigle week in Supplementary file, for the lack of space.

**Table 1:**
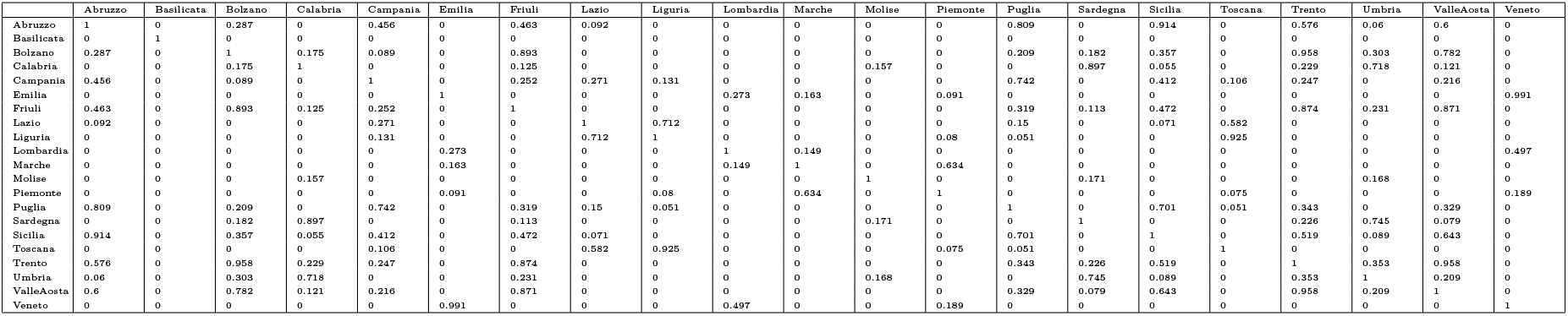
Similarity Matrix of Hospitalised with Symptoms network

Results show that according the type of data, a significant difference exists (p-value less than 0.05) among some regions while for others it is possible to highlight statistically similar distributions. Also, the significance varies by performing the analysis on whole selected time interval and on single week.

Finally, we used Kruskal-Wallis test performing an analysis on the same type of data for all regions (i.e. carrying out multiple comparisons) for the observation period and then, for each single week. Kruskal-Wallis test is a non-parametric method for analysis of variance used to determine if more samples originate from the same distribution. The results confirmed a significant difference considering all regions on the same type of data for the observation period for each single week.

### 2.3 Network-based Analysis

In order to evaluate the evolution of Italian COVID-19 data and evidence which regions show similar trend, we built networks of each data [1] starting from the result of Wilcoxon test. The nodes of the networks are the Italian regions and the edges link two regions (nodes) with similar trend according to significance level (p-value *>* 0.05) obtained from Wilcoxon test, otherwise (p-value *<* 0.05) there is not connection among nodes. The length of the edges is inversely proportional to nodes similarity. The network analysis is performed using the igraph libraries [3].

At first, we built ten networks, one for each data (Hospitalised with Symptoms, Intensive Care data, Total Hospitalised, Home Isolation, Total Currently Positive, New Currently Positive, Discharged/Healed, Deceased, Total Cases, Swab) by considering the period February 24 to March 29. Then, we build the same networks by considering single weeks. The ten networks for the observation period and for the five weeks are reported in Figures 2-11.

**Figure 2:**
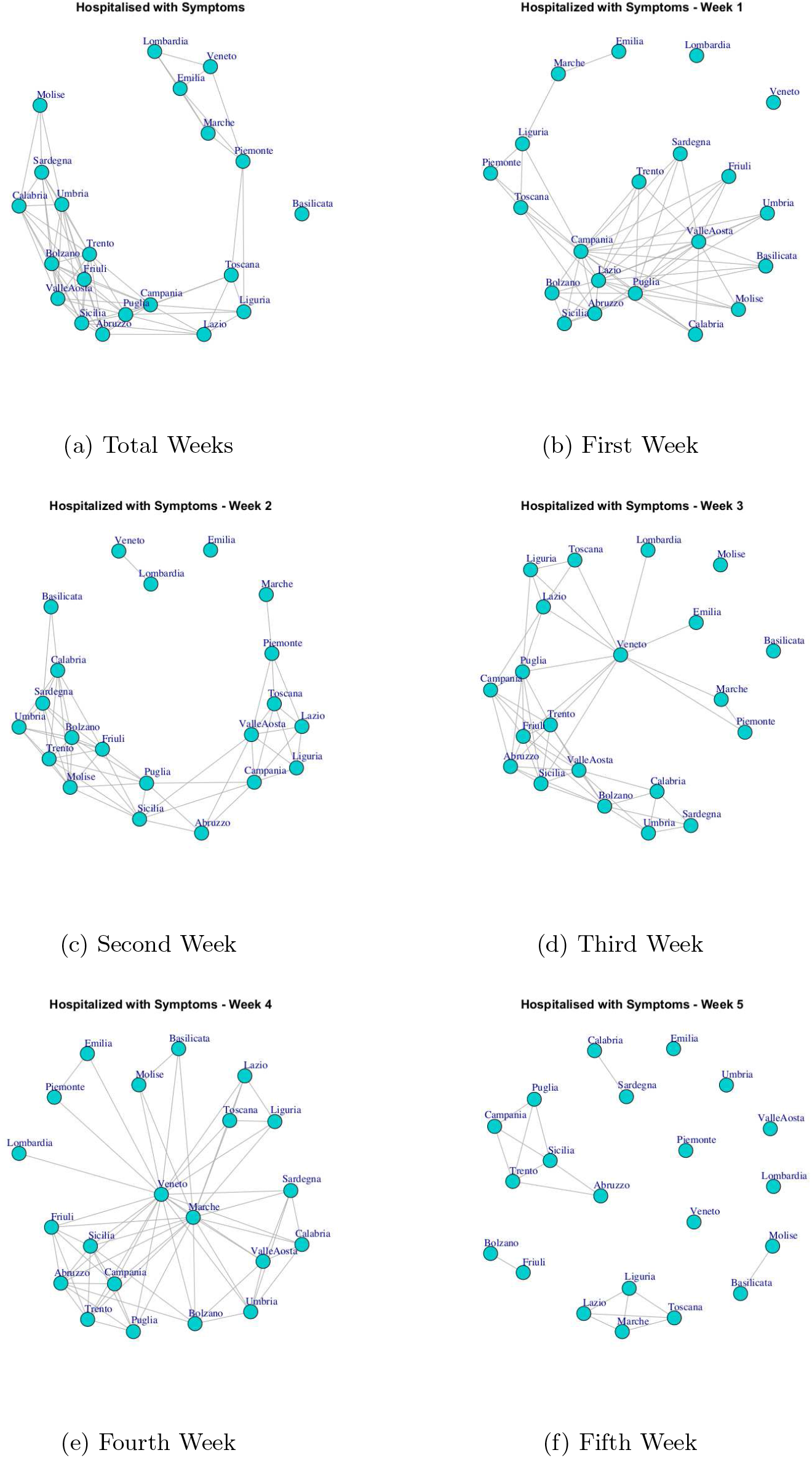
Evolution of Hospitalised with Symptoms Network

Then, starting from the ten networks related to all five weeks, we wanted to identify which regions form a community from the similarity point of view. For this, we applied Walktrap community finding algorithm [7] that identifies densely connected subgraphs, i.e communities, in a graph via random walks. The idea is that short random walks tend to stay in the same community.

The extracted communities from all Italian COVID-19 networks are reported in Figure 12.

Finally, we analize the evolution of the extracted communities over a period of time. We want to evaluate how different data present different communities and how the communities are different considering different temporal interval on the same data. We analized the evolution of the communities at different time points, i.e. at the end of the first week, after tree weeks and after five weeks (the observation period). The Figure 13-22 report the evolution of the communities related to different data.

## 3 Results and Discussion

### 3.1 Temporal Evolution of Networks

In this section, we analyze the evolution of the Italian COVID-19 networks data. Figure 2 shows the evolution of Hospitalised with Symptoms network over five weeks. Figure 2(a) reports the network that represents the behaviour of regions respect to number of hospitalised patients up to March 29, whereas Figure 2(b), (c), (d), (e), (f) represent the networks on each single week. It is to notice, the network structure changes according to the analyzed time interval. At the end of 35th day, the network has all nodes connected with exception of a single node that represent the Basilicata region. Furthermore, it is possibile to highlight different community structures consisting of groups of regions with similar trends. Figure 12(a) reports five communities: the first one consists of Basilicata; the second one is composed by Piemonte, Marche, Emilia, Lombardia, Veneto; the third one is composed by Liguria, Lazio, Toscana; the fourth one is formed by Campania, Puglia, Sicilia, Abruzzo, Valle d’ Aosta, Friuli, Trento, Bolzano; the last one is composed by Umbria, Sardegna, Calabria, Molise.

Figure 3 represents the evolution of Intensive Care network. It is possible to notice that Lombardia and Veneto, that are the most affected region by Coronavirus disease with high number hospitalised in Intensive Care Units, are disconnected in the first week. In the second week, Lombardia and Veneto are connected by an edge that represents a level of similarity, whereas in the fifth week Veneto is linked with Emilia and Lombardia and this group of regions becomes a disconnected component among with other regions. Thus, while initially Lombardia and Veneto showed a similar trend of Intensive Care data, after Veneto moved far from Lombardia trend. Furthermore, by analyzing the communities in the Intensive Care network (Figure 12(b)), it is possible to evidence four subgraphs formed by (i) Lombardia and Veneto, (ii) Umbria and Lazio, (iii) Marche, Emilia, Piemonte, Toscana and (iv) a large module formed by Campania, Sicilia, Sardegna, Abruzzo, Umbria, Calabria, Basilicata, Bolzano, Valle d’Aosta, Friuli, Trento, Molise.

**Figure 3:**
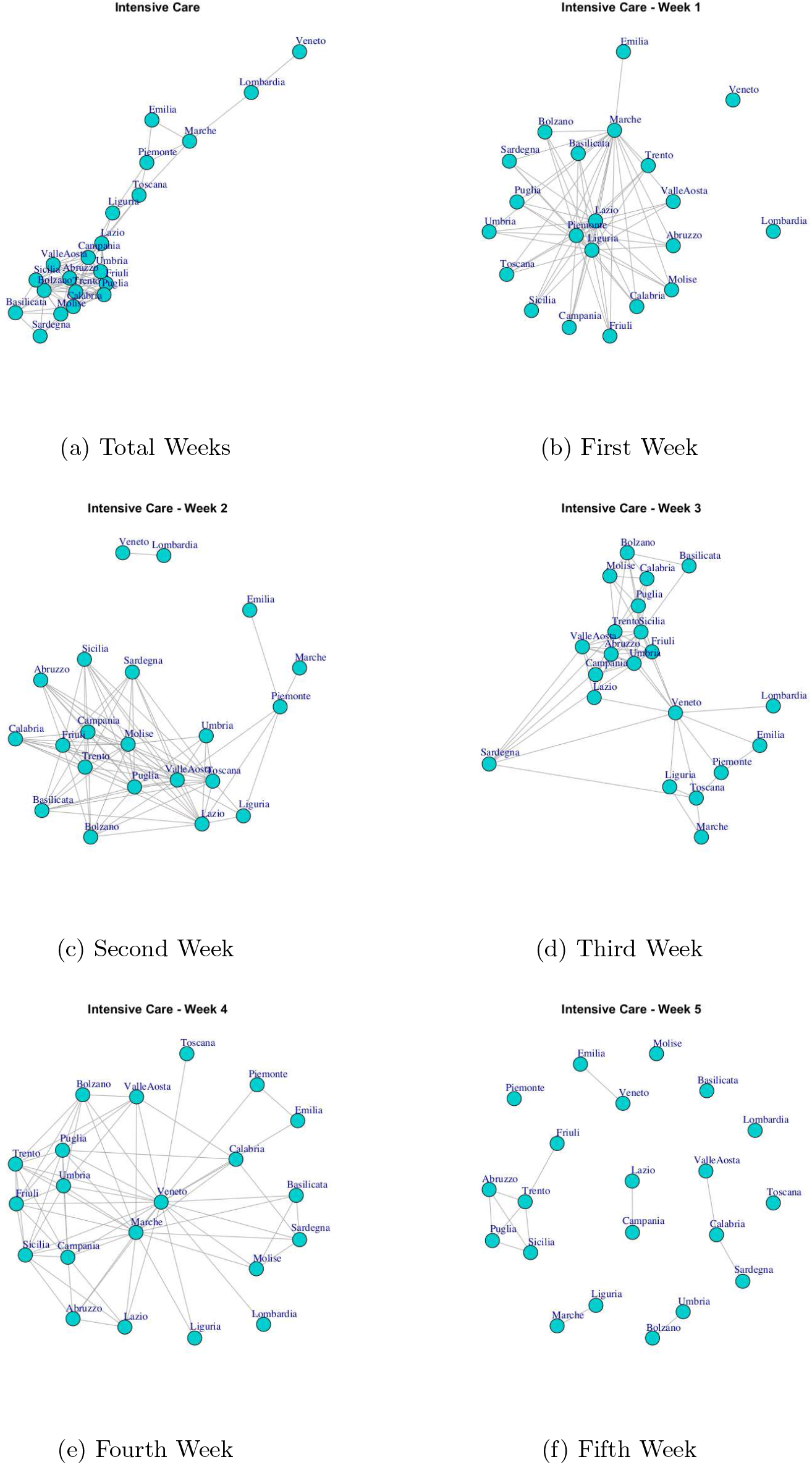
Evolution of Intensive Care Network

Figure 4 show the Total Hospitalised network evolution. Starting form the first week, the structure of the network has two disconnected nodes representing Lombardia and Veneto, whereas in the second week the network evolves by presenting a single disconnected node that represent Emilia, and two connected nodes, Lombardia and Veneto, which in turn are disconnected from dense subgraph. In the third and fourth week the network structure presents all connected components, and high number of nodes result disconnected in the fifth week. Finally, all regions are connected in the final network. By analyzing the communities detected in Total Hospitalised network (Figure 12(c)), it is possible to notice a similarity with those extracted by Hospitalised with Symptoms networks. In fact, there is a correspondence among three communities: (i) Basilicata and Piemonte, (ii) Marche, Emilia, Lombardia, (iii) Veneto and Liguria, Lazio, Toscana. This means that those regions that form the tree communities present the same behaviour according to the number of the hospitalised patients with symptoms and the total number of hospitalised patients.

**Figure 4:**
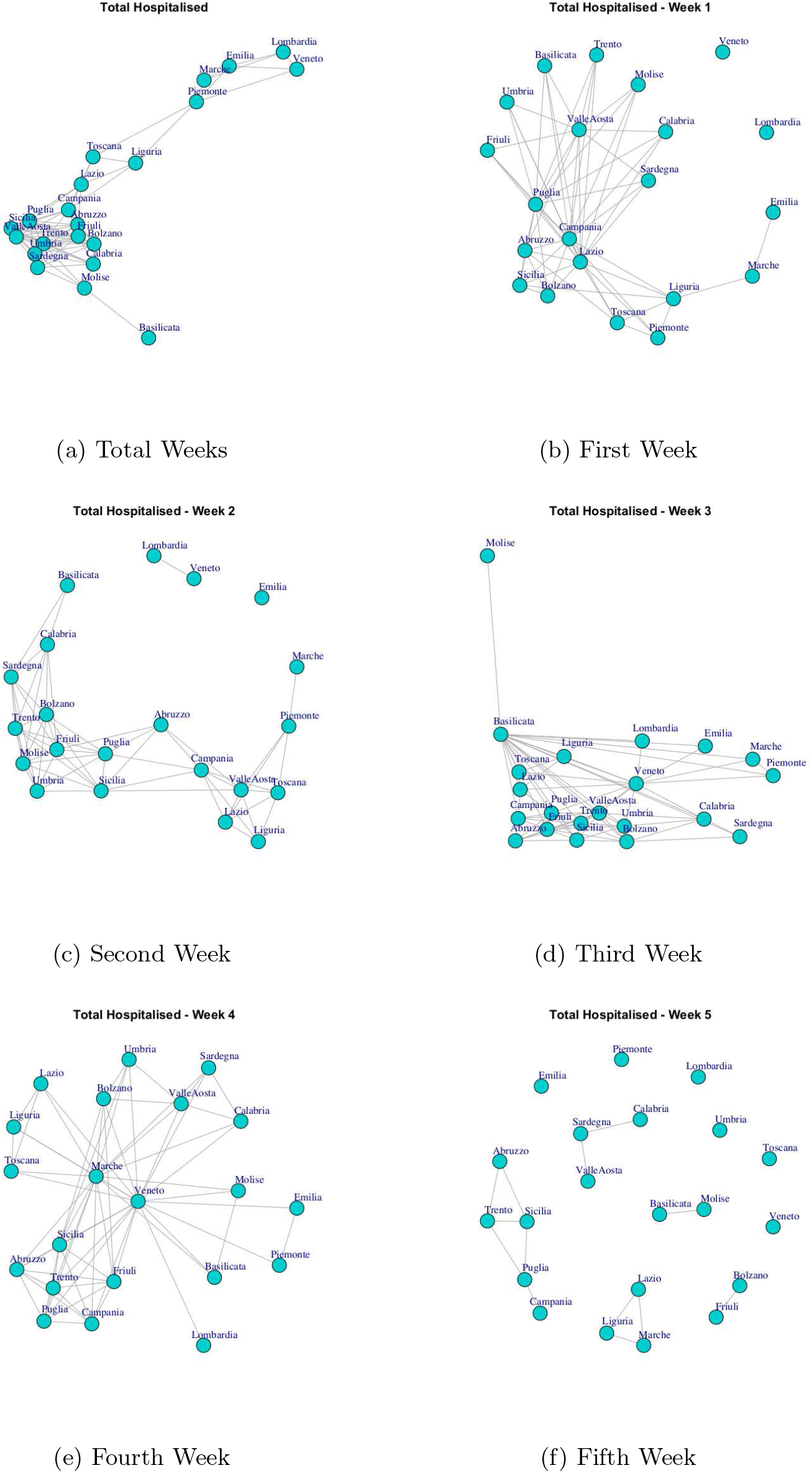
Evolution of Total Hospitalised Network

Figure 5 shows the evolution of Home Isolation network over five weeks. In first week, Lombardia, Veneto and Emilia show a different trend of the number of subjects in isolation at home, both them and also compared to other regions. Next, in the network resulting by considering the all weeks, Lombardia, Veneto, Emilia and Marche formed a subgraph disconnected by a different dense subgraph composed by the rest of the regions. However, Veneto represents a single community in Figure 12
(d). This means that the behaviour of Veneto presents a low similarity respect to Lombardia, Emilia and Marche despite forming a module.

**Figure 5:**
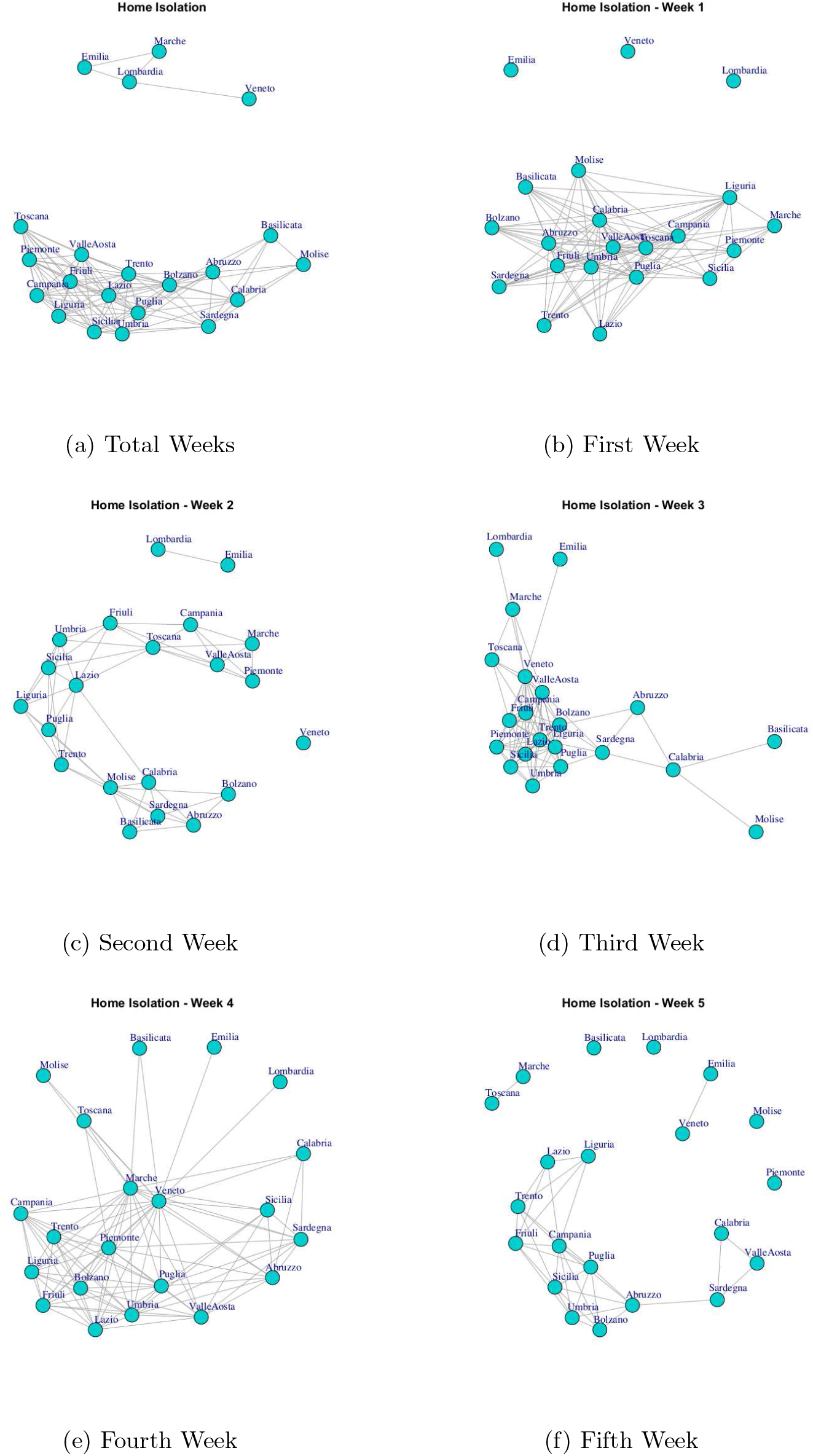
Evolution of Home Isolation Network

Figure 6 and Figure 7 show the Total Currently Positive network and New Currently Positive network. Both networks evolve over five weeks by forming a structure in which all nodes are connected. According to the extracted communities, four modules are identified in Total Currently Positive network (see Figure 12(e)) and they are formed by: (i) Piemonte; (ii) Lombardia, Veneto, Marche, Emilia, (iii) Basilicata, Molise, Calabria, Sardegna, (iv) Puglia, Friuli, Valle d’ Aosta, Toscana, Lazio, Abruzzo, Umbria, Campania, Trento, Liguria Bolzano, Sicilia. The communities identified in New Currently Positive network in Figure 12(f) are: (i) Piemonte, Marche, Toscana; (ii) Lombardia, Veneto, Emilia, (iii) Basilicata, Molise, (iv) Puglia, Friuli, Valle d’ Aosta, Lazio, Abruzzo, Umbria, Campania, Trento, Liguria Bolzano, Calabria, Sardegna, Sicilia.

**Figure 6:**
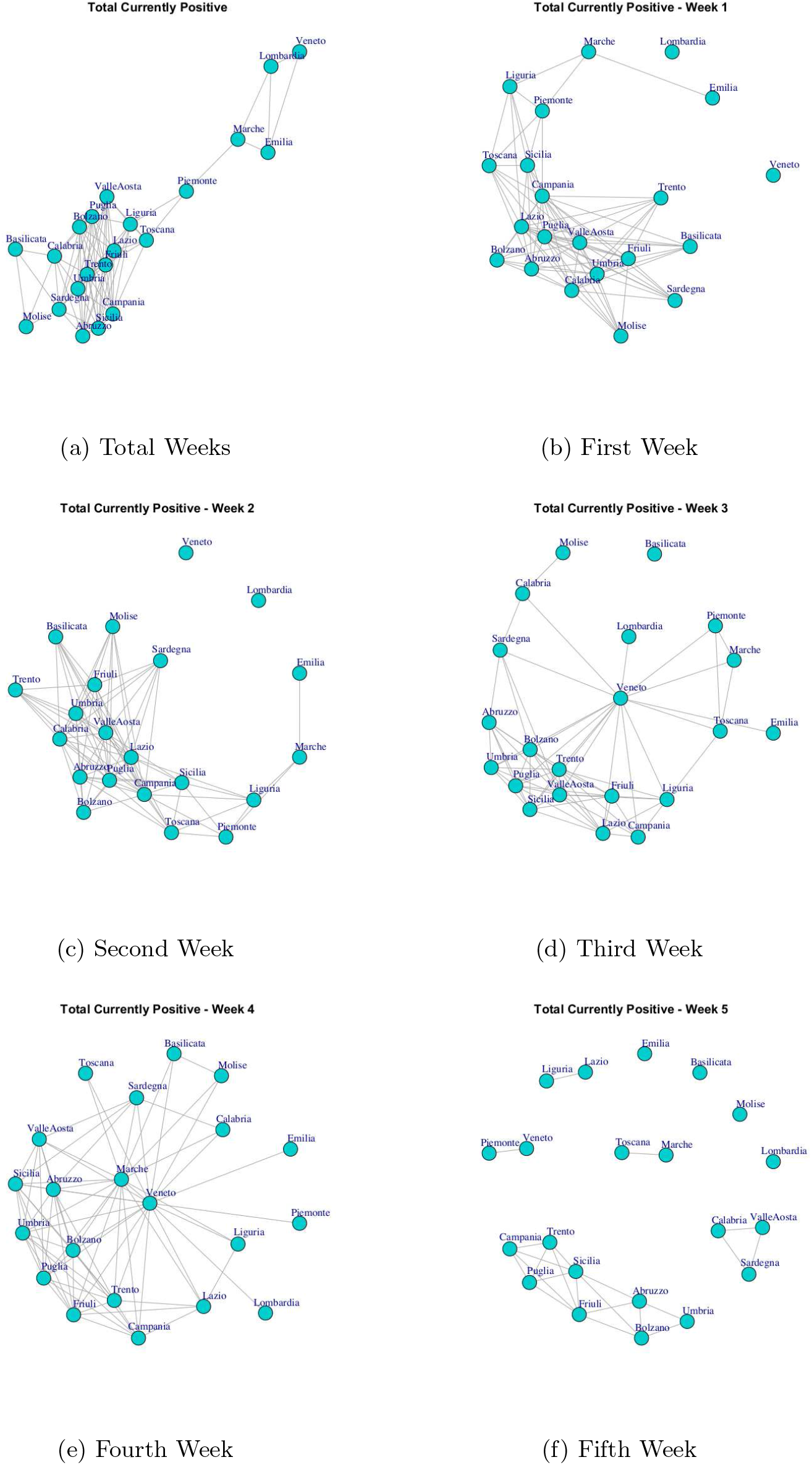
Evolution of Total Currently Positive Network

**Figure 7:**
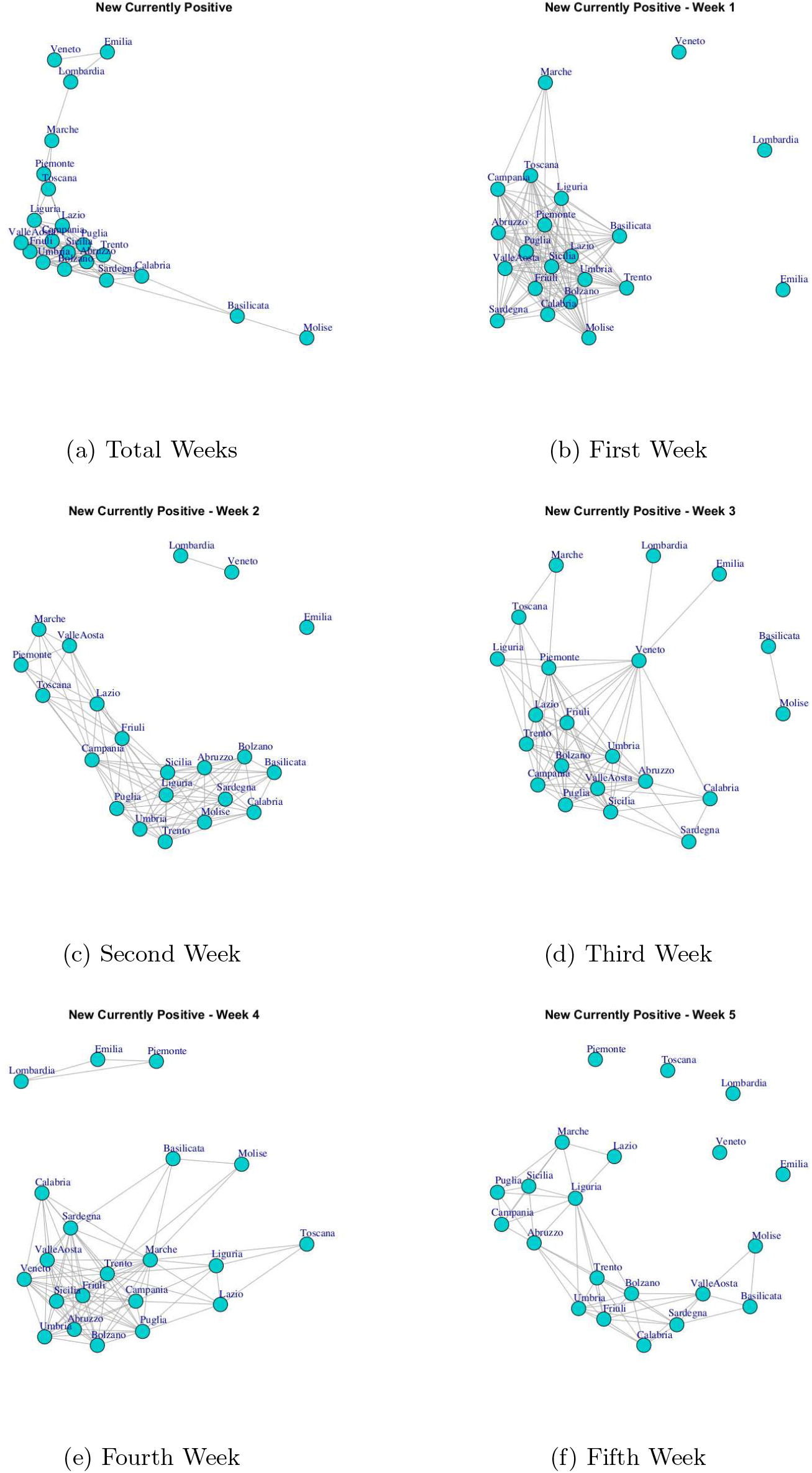
Evolution of New Currently Positive Network

It is possible to notice that: Lombardia, Veneto, Emilia form a community in both Total Currently Positive network and New Currently Positive network, Piemonte represent in sigle community in Total Currently Positive network and while Piemonte is associated with Marche and Toscana in New Currently Positive network.

Figure 8 represents the Discharged/ Healed network over the five weeks. In the first week, the network structure present all nodes connected with exception of Veneto. In the second week, Discharged/ Healed network is formed by three subgraphs; in the third and fourth weeks the network results very dense; in the fifth week is caracterized by different disconnected components, and finally, at the end of 35 days the network is composed by a subgraph composed by Lombardia and Veneto and another subgraph highly connected. This means that Lombardia and Veneto have a similar behaviour that it is different by the rest of the Italian regions. Also Lombardia and Veneto represents one of the five communities extracted by Discharged/ Healed network. The extracted communities are reported in Figure 12(g).

**Figure 8:**
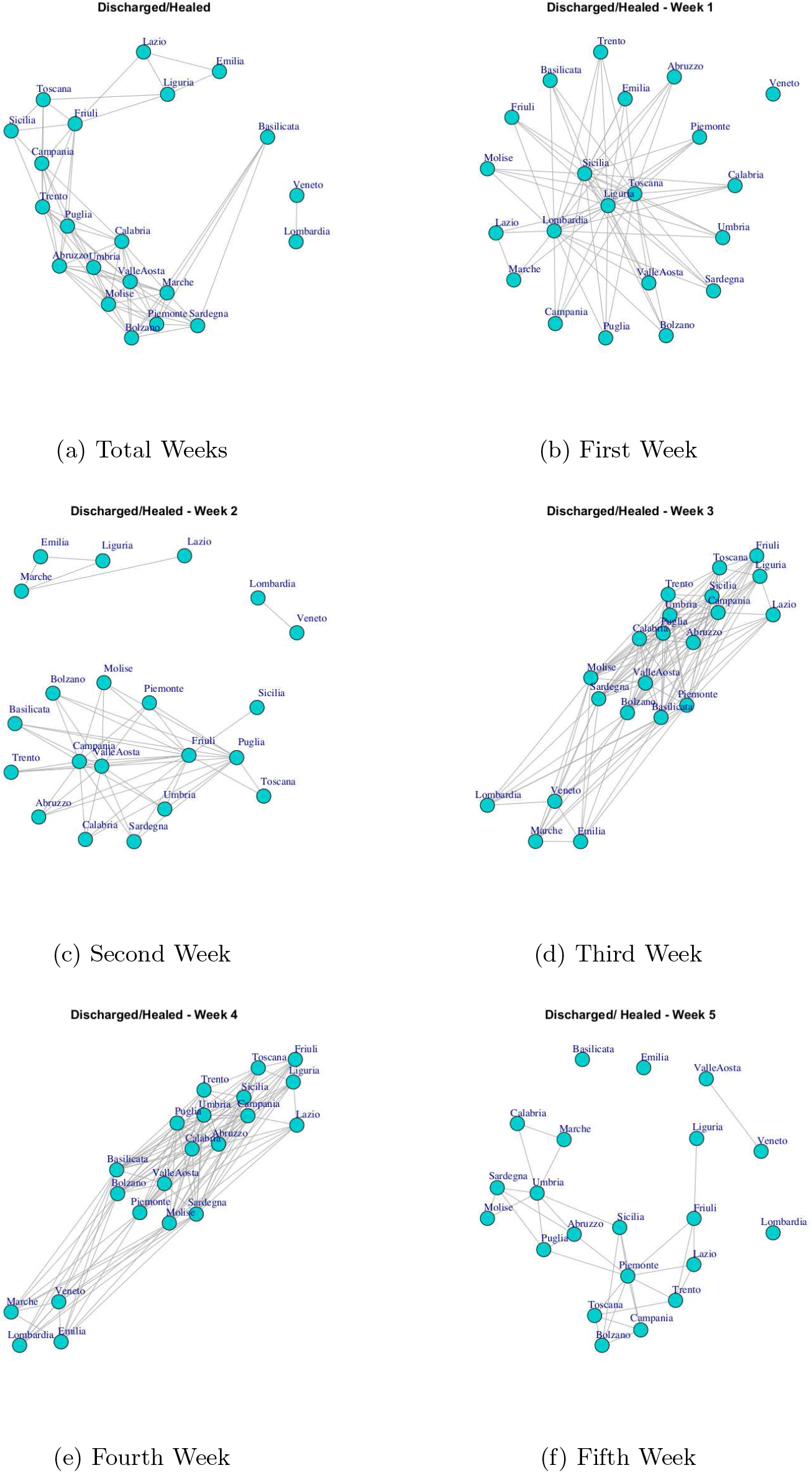
Evolution of Discharged/ Healed Network

Figure 9 shows the evolution of Deceased network. The evolution of this network is different by other Italian COVID-19 networks. In fact, in first week all nodes are disconnected, so all Italian regions present different trend. In the second week, it is possible to notice that Emilia and Marche nodes are disconnected and there is a subgraph composed by Lombardia and Veneto and then there is a large subgraph formed by other regions. In the third week all nodes present connections. In the fourth and and fifth week the Deceased network presents different disconnected components, then, the final network shows a single disconnected node that represents Basilicata. Also the Basilicata represents a single community of Deceased network, see Figure 12(h). The others extracted communities are: (i) Piemonte, Toscana Liguria, Lazio, Friuli, Puglia, Valle d’ Aosta, (ii) Lombardia, Veneto, Emilia, Marche, (iii) Sicilia, Molise, Abruzzo, Umbria, Campania, Trento, Bolzano, Calabria, Sardegna.

**Figure 9:**
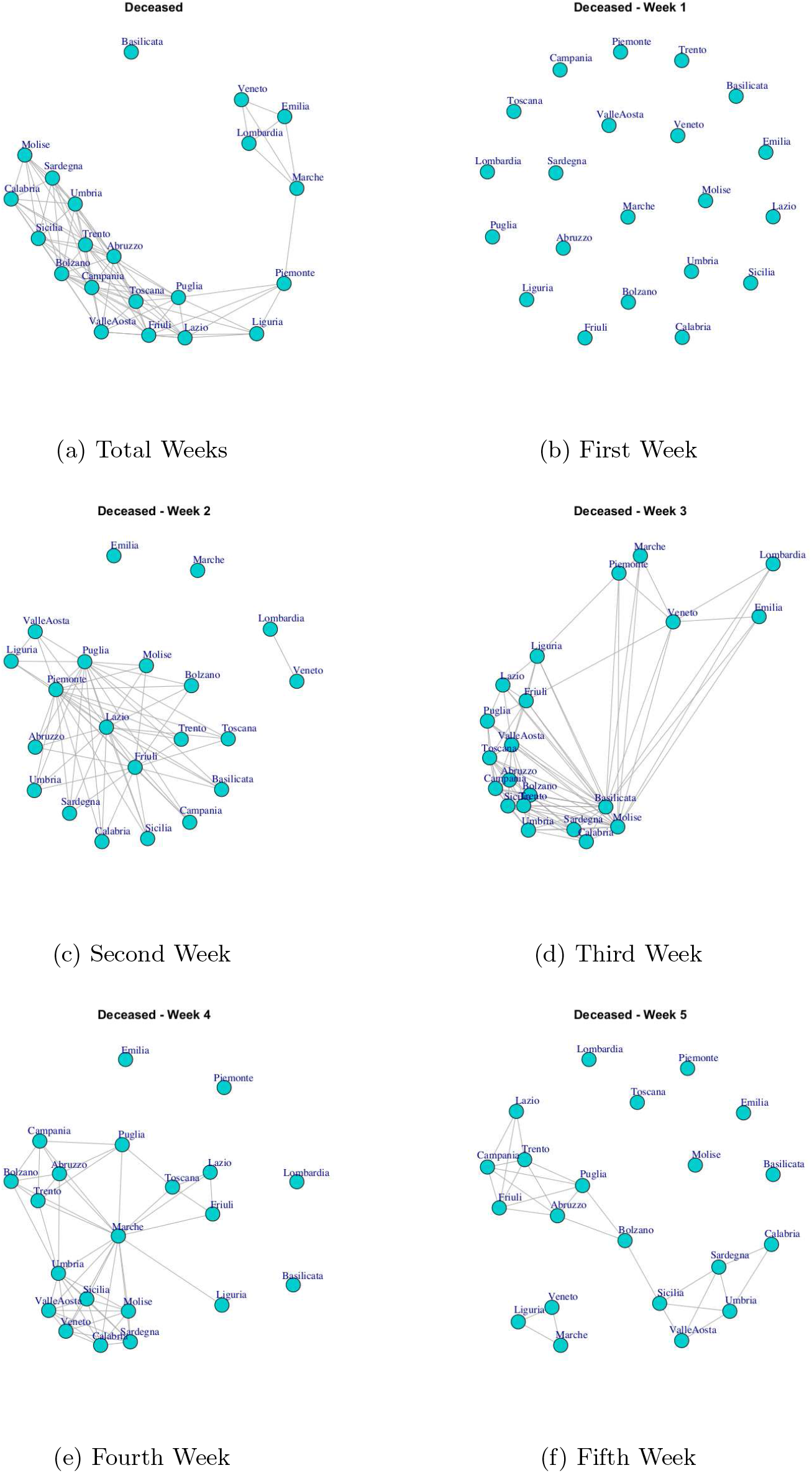
Evolution of Deceased Network

Figure 10 represents the Total Cases network over the five weeks. The final network evidences that the Italian regions present a significant level of similarity respect to the number of total Coronavirus cases because all nodes are connected. Figure 12(i) show the communities identified in Total Cases. The first community is composed by Lombardia, Veneto, Emilia, Marche; the second community is composed by Piemonte; the third community is composed by Basilicata, Molise; the fourth community is composed by Toscana Liguria, Lazio, Campania, Friuli, Sicilia Puglia, Valle d’ Aosta formed, whereas Abruzzo, Umbria, Trento, Bolzano, Calabria, Sardegna formed the fifth community.

**Figure 10:**
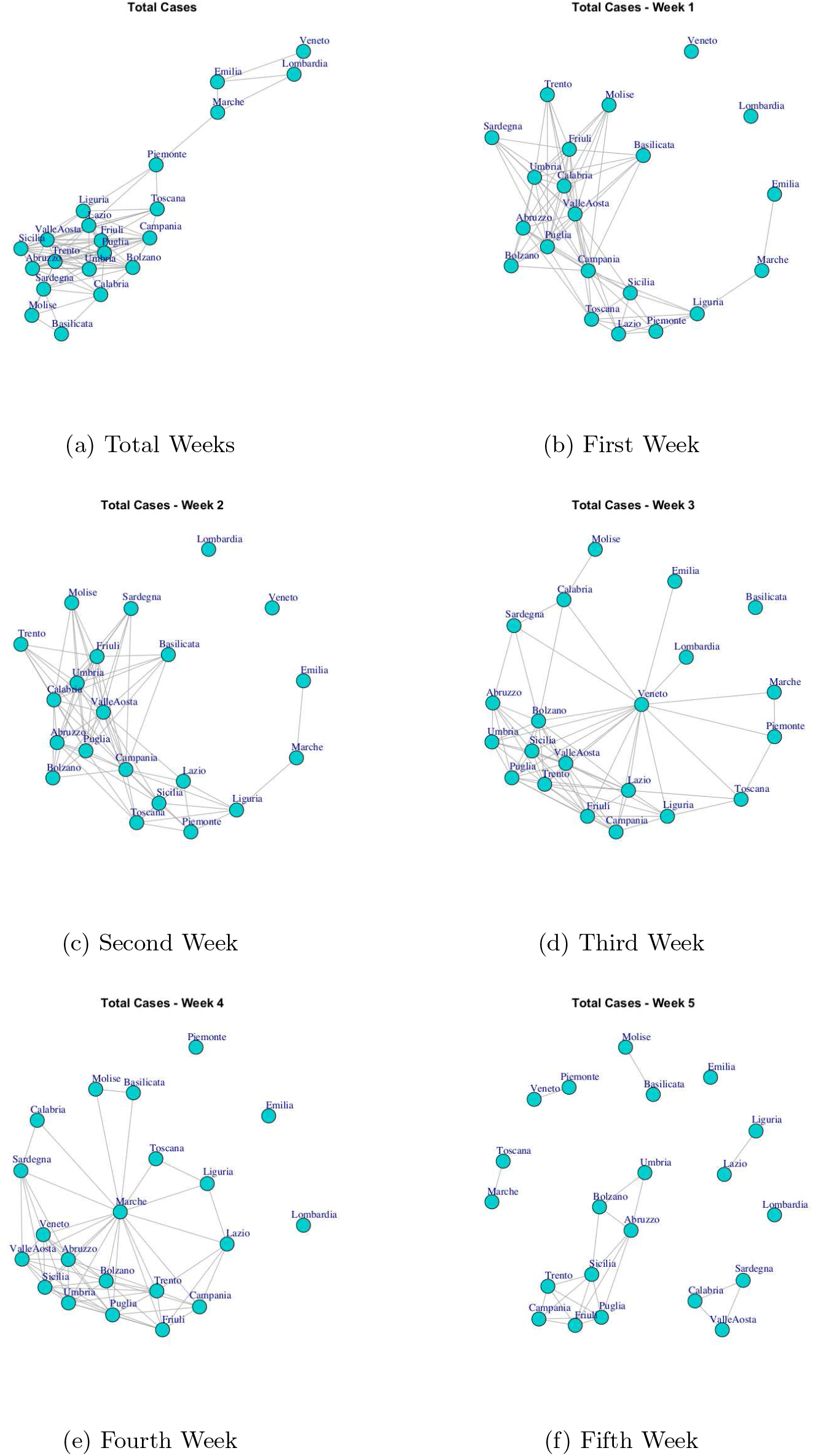
Evolution of Total Cases Network

Figure 11 shows the evolution of Swab Network that represents the number of performed swab tests. The network, in the first week, shows Lombardia and Veneto nodes disconnected by other regions. In fact they are the Italian regions that initially performed high number of test swabs. Also, Veneto region has not connections in the final network and this reflects the policy of the Veneto to carry out swab tests on asymptomatic subjects, i.e. it is an outlier with respect to other regions. Figure 12(j) shows the extracted communities in the Swab network. The first community is composed by Veneto; the second community is composed by Lombardia, Emilia; the third community is composed by Basilicata, Molise; the fourth community is formed by Marche, Toscana, Lazio, Piemonte, Friuli, Valle d’Aosta; the fifth community is formed by Sicilia, Campania, Liguria, Puglia; while Abruzzo, Umbria, Trento, Bolzano, Calabria, Sardegna formed the sixth community.

**Figure 11:**
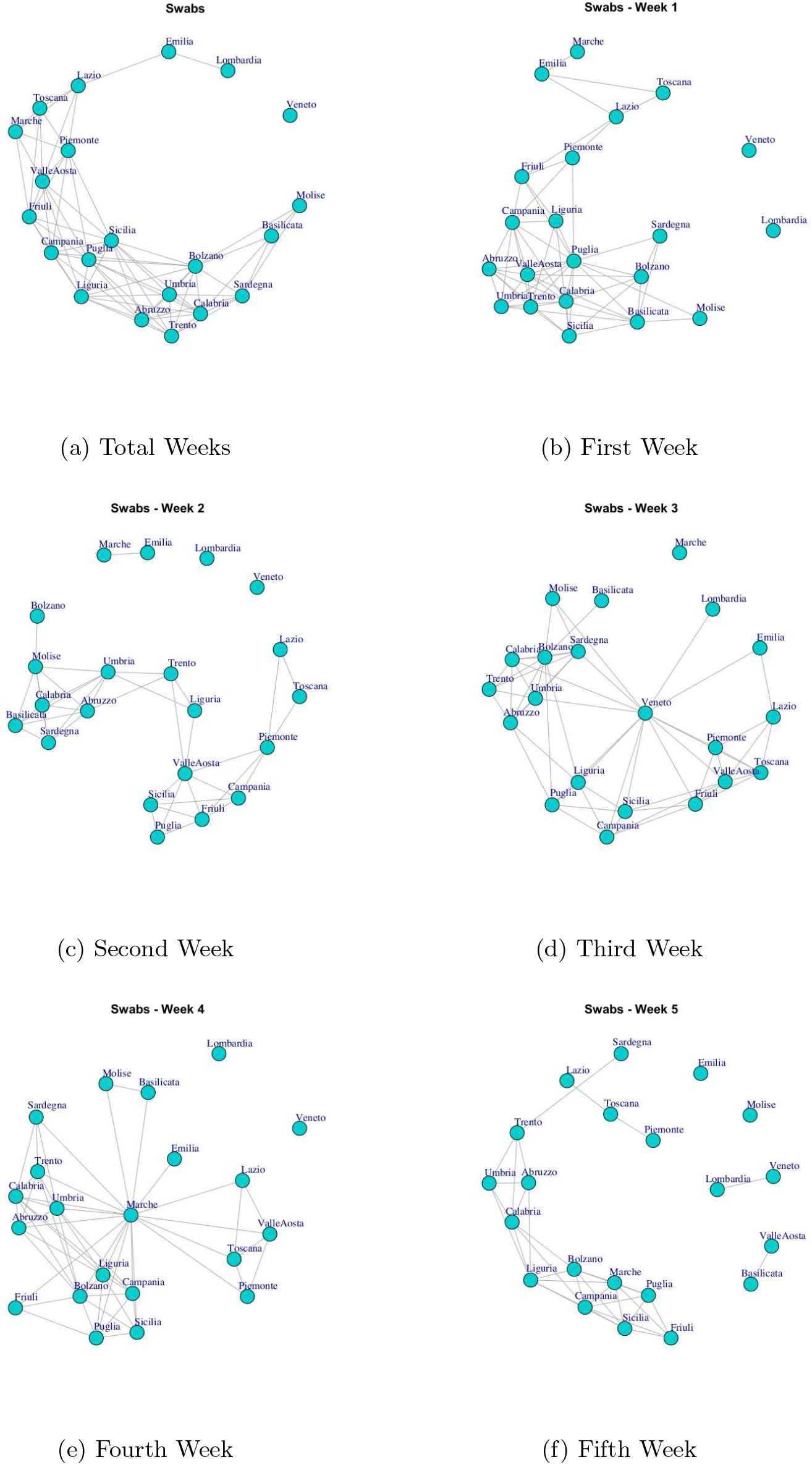
Evolution of Swabs Network

### 3.2 Analysis of Communities

In this section, we analize the evolution of communities over a period of time. We consider three different temporal zoom: at the end of the fist week, after three weeks and in the observation period.

Figure 13 reports the evolution of Hospitalised with Symptoms Network Communities.

Figure 13(a) reports six communities extracted in the first week: (i) Lombardia, (ii) Veneto, (iii) Emilia, Marche, (iv)Liguria, Toscana, Piemonte, (v)Puglia, Lazio, Campania, Abruzzo, Bolzano, Sicilia, (vi) Umbria, Sardegna, Calabria, Molise, Valle d’Aosta, Friuli, Basilicata, Trento.

At the end of three weeks, Venezia that represents a community in the previous week, moves in another community, whereas Emilia leaves the community with Marche and becomes a single community, also some regions migrate from fifth and sixth communities to other communities. So, Figure 13(b) reports the five extracted communities after three weeks: (i) Lombardia, (ii)Emilia, (iii)Veneto, Marche, Piemonte, Liguria, Toscana, Lazio, (iv) Trento, Bolzano, Abruzzo, Friuli, Sicilia, Puglia, (v)Campania, Umbria, Sardegna, Calabria, Molise, Valle d’Aosta, Basilicata. Finally, Figure 13(c) reports five communities in the observation period: the first one consists of Basilicata that leaves the previous community and becomes a single one; the second one is composed by Piemonte, Marche, Emilia, Lombardia, Veneto; the third one is composed by Liguria, Lazio, Toscana; the fourth one is formed by Campania, Puglia, Sicilia, Abruzzo, Valle d’Aosta, Friuli, Trento, Bolzano; the last one is composed by Umbria, Sardegna, Calabria, Molise.

Figure 14 reports the evolution of Intensive Care Network Communities. It is possible to notice that in the first week, Figure 14(a), there are: a large community formed by Umbria, Lazio, Piemonte, Toscana Campania, Sicilia, Sardegna, Abruzzo, Umbria, Calabria, Basilicata, Bolzano, Valle d’Aosta, Friuli, Trento, Molise; two single communities formed by Lombardia and Emilia; and a small community formed by Emilia and Marche.

**Figure 12:**
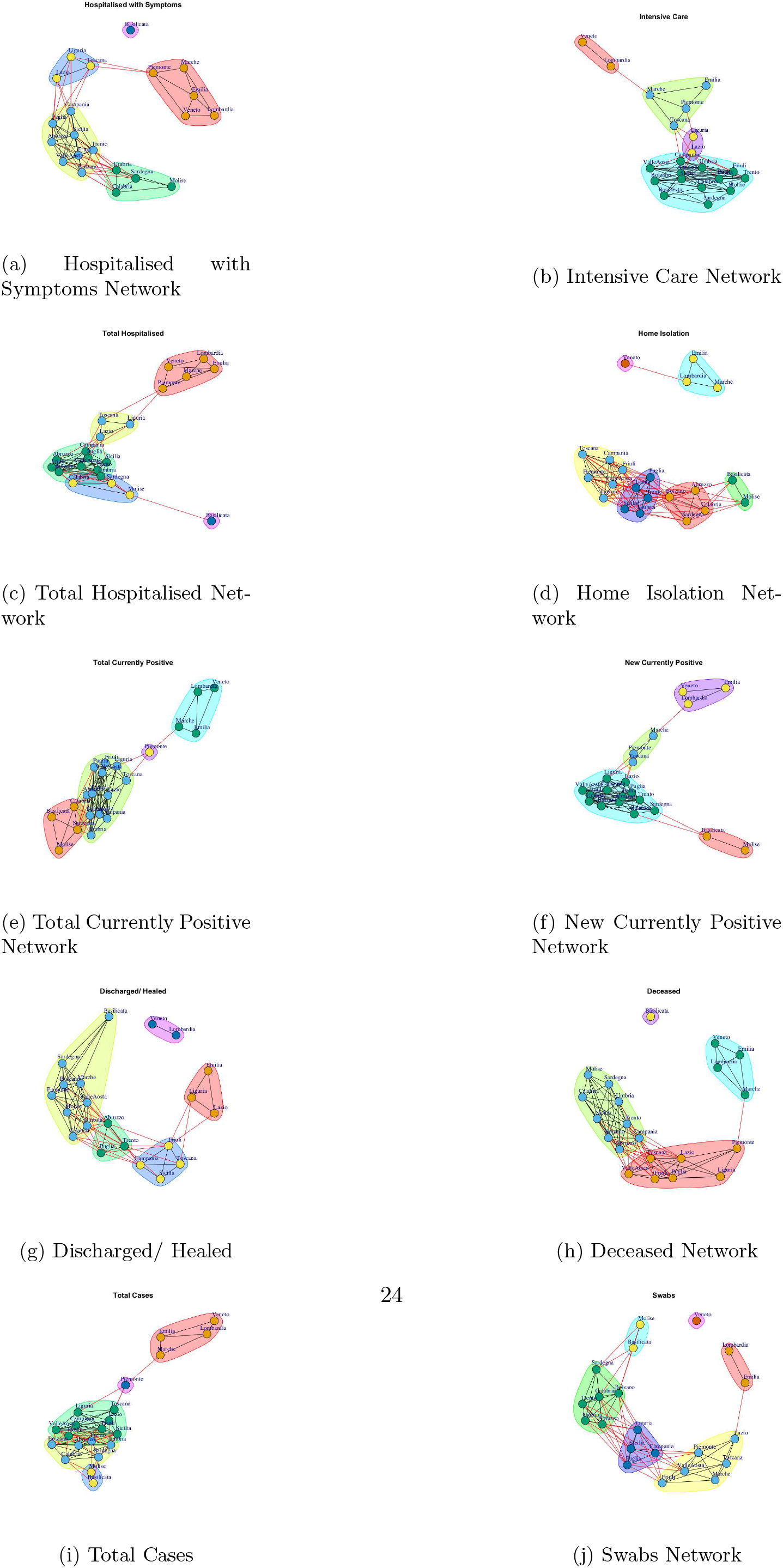
The figure show the communities identified in Italian COVID-19 networks.

**Figure 13:**
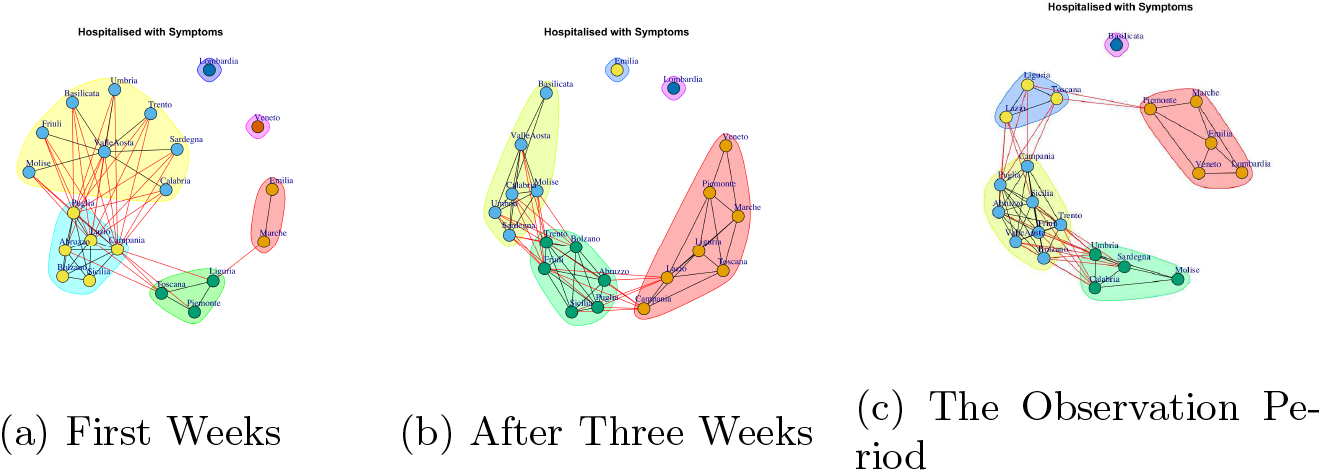
Evolution of Hospitalised with Symptoms Network Communities.

**Figure 14:**
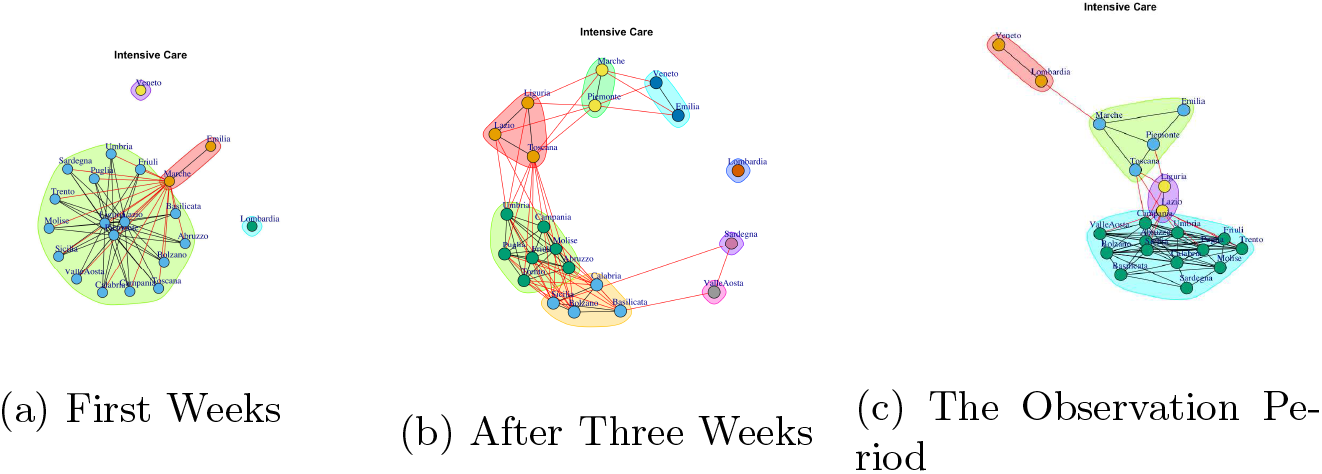
Evolution of Intensive Care Network Communities.

After three weeks the number of extracted communities increases, see Figure 14(b). In fact, Lombardia, Sardegna, Valle d’Aosta represent three single communities, Veneto and Emilia form a community, as well as, Marche and Piemonte. Then, Liguria, Lazio and Toscana form a six community, and the last two are composed by (i) Umbria, Campania, Molise, Abruzzo, Friuli, Trento, Puglia and (ii) Calabria, Sicilia, Bolzano, Basilicata.

Finally, in the observation period, five communities are mined, see Figure 14(c), formed by (i) Lombardia and Veneto, (ii) Liguria and Lazio, (iii) Marche, Emilia, Piemonte, Toscana and (iv) a large module formed by Campania, Sicilia, Sardegna, Abruzzo, Umbria, Calabria, Basilicata, Bolzano, Valle d’Aosta, Friuli, Trento, Molise.

Figure 15 reports the evolution of Total Hospitalised Network Communities.

**Figure 15:**
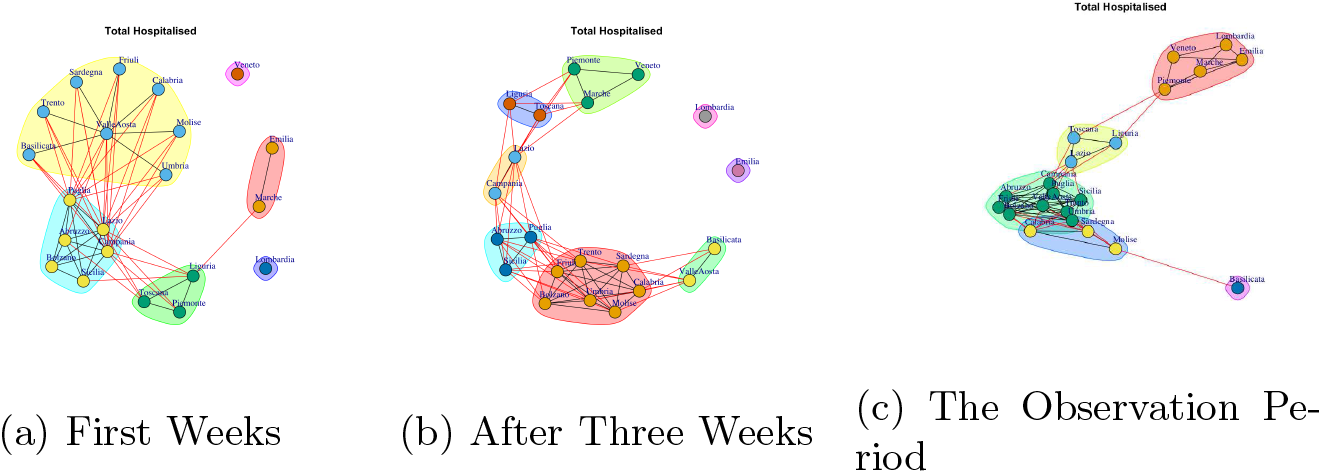
Evolution of Total Hospitalised Network Communities.

Figure 15(a) shows the five mined communities. The fist community is composed by Liguria, Toscana, Piemonte, the second one is formed Sardegna, Umbria, Calabria, Basilicata, Valle d’Aosta, Friuli, Trento, Molise; the third module comprises Lazio, Campania, Sicilia, Abruzzo, Bolzano, Puglia; the forth community is represented by Marche, Emilia; the fifth is represented by Lombardia and the last one consists of Veneto.

After three weeks (see Figure 15(b)) the regions moves,, with the exception of Lombardia that continues to represent a single community and the communities becomes eight. In fact, Emilia becomes a single community; Veneto becomes a community among with Piemonte and Marche; Toscana moves in the community with Liguria; Lazio and Campania forms a new community, as well as, Basilicata and Valle d’Aosta; another community is formed by: Abruzzo, Puglia, Sicilia and the last one is formed by Friuli, Bolzano, Trento, Umbria, Sardegna, Calabria, Molise.

At the end of the observation period the five communities, reported in Figure 15(b), are formed. The first one consists of Basilicata that leaves the previous community and becomes a single one; the second one is composed by Toscana, Liguria and Lazio; Sardegna, Calabria, Molise leaves the previous large community and form a smaller one; by the fourth one is formed by Piemonte, Marche, Emilia, Lombardia, Veneto; the last one is composed by Umbria, Puglia, Sicilia, Abruzzo, Valle d’Aosta, Friuli, Trento, Bolzano;

Figure 15 reports the evolution of Home Isolation Network Communities.

Figure 16(a) reports the mined communities at the end of first week. It is possible to notice that Lombardia, Veneto and Emilia form single communities, and then there are three large communities: the first one is represented by Piemonte, Liguria, Marche, Sicilia, Campania; the second one is composed by Puglia, Valle d’Aosta, Toscana, Umbria, Calabria; the third one Trento, Lazio, Abruzzo, Sardegna, Bolzano, Basilicata and Molise, Friuli, Campania.

**Figure 16:**
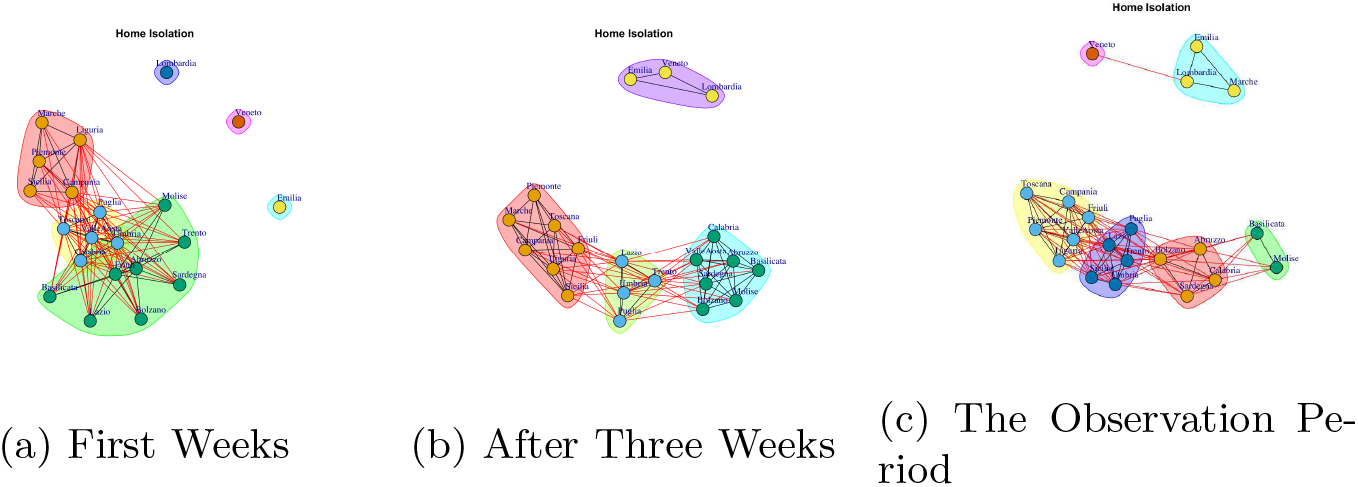
Evolution of Home Isolation Network Communities.

At the end of three week, Lombardia, Veneto and Emilia move together to form a unique community, whereas, the other regions form new communities, such as, (i) Puglia, Trento, Lazio, Umbria, (ii) Calabria, Abruzzo, Valle d’Aosta, Sardegna, Bolzano, Basilicata and Molise, (iii) Sicilia, Toscana, Friuli, Piemonte, Liguria, Marche, Campania (see Figure 16(b)).

Figure 16(c) shows the communities topology in the observation period. Veneto leaves the community among with Lombardia and Emilia and it becomes a single one, whereas, Lombardia, Emilia forms a new module among with Marche. Basilicata and Molise move together to form a unique community. Sardegna, Calabria, Abruzzo, Bolzano form a forth community. The fifth community is composed by Puglia, Trento, Lazio, Umbria, Sicilia, and the sixth one is represented by Toscana, Piemonte, Valle d’Aosta, Friuli, Liguria, Campania.

Figure 17 reports the evolution of Total Currently Positive Communities. At the end of first week the mined communities are eight and they are reported in Figure 17(a). The fist community is composed by Umbria, Sardegna, Basilicata, Molise, Friuli Toscana, Calabria, Valle d’Aosta, Trento,; the second one is formed by Bolzano, Lazio, Abruzzo, Puglia; the third module comprises Campania, Sicilia Liguria; Piemonte and Lazio represent the forth community; the fifth one consists of Marche; the sixth community is represented by Emilia; the seventh is represented by Lombardia and the last one consists of Veneto.

**Figure 17:**
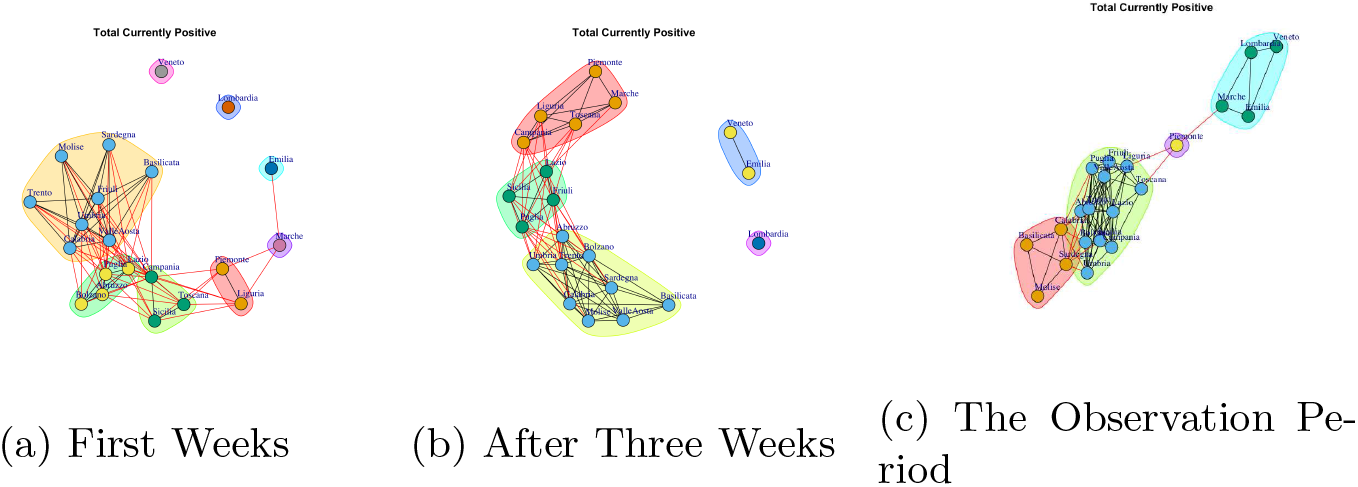
Evolution of Total Currently Positive Network Communities.

After three weeks, the number of communities (see Figure 17(b)) decreases. In fact, it is possible to notice five subgraphs. Emilia joints with Veneto and Lombardia remains single community. Lazio, Sicilia, Friuli, Puglia form a new community; Piemonte, Toscana, Campania, Marche, Liguria, represent a forth community; Trento, Abruzzo, Umbria, Calabria, Sardegna, Basilicata, Molise, Bolzano, Campania, Valle d’Aosta, form a fifth community;

In the observation period, the number of extracted communities further decreases, see Figure 17(c) The first community is composed by Veneto, Lombardia, Emilia, Marche; the second community is composed by Piemonte; the third community is composed by Basilicata, Molise, Calabria, Sardegna; the fourth community is formed by Toscana, Lazio, Friuli, Valle d’ Aosta, Sicilia, Campania, Liguria Puglia, Abruzzo, Umbria, Trento, Bolzano.

Figure 18 reports the evolution of New Currently Positive Communities. Figure 18(a) reports the mined communities at the end of first week. It is possible to notice that Lombardia, Veneto and Emilia form single communities, and then there are two large communities: the first one is represented by Marche, Piemonte, Liguria, Campania, Abruzzo, Toscana; the second one is composed by Puglia, Valle d’Aosta, Umbria, Calabria, Sicilia, Campania, Trento, Lazio, Sardegna, Bolzano, Basilicata and Molise, Friuli.

**Figure 18:**
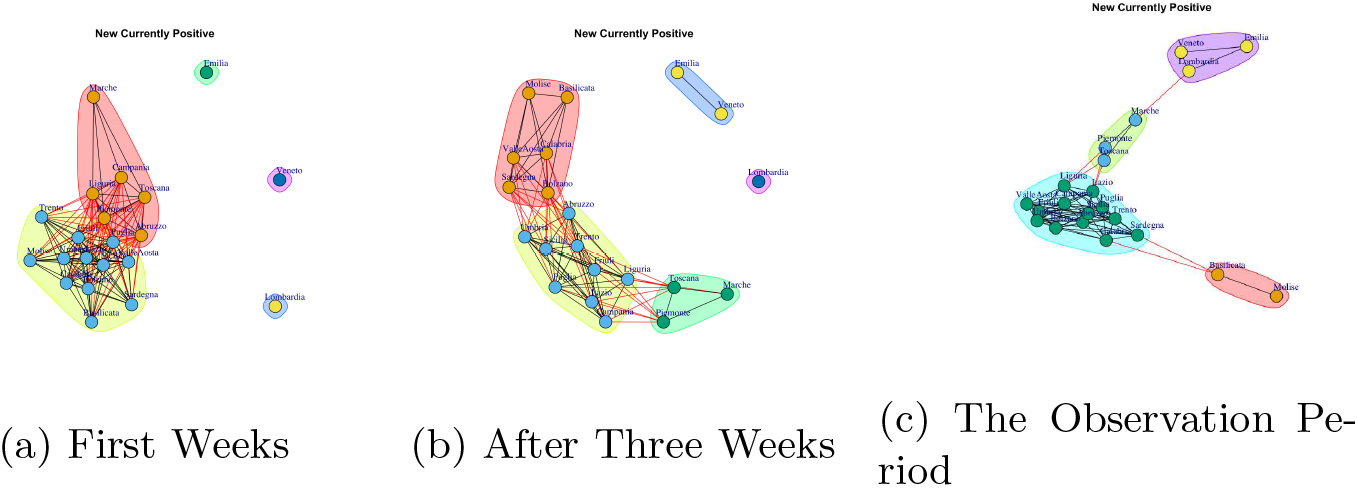
Evolution of New Currently Positive Network Communities.

After three weeks, the extracted communities remain five but the regions that form them vary, see Figure 18(b). The first community is composed by Lombardia; the second community is composed by Veneto and Emilia; the third community is composed by Basilicata, Molise, Valle d’ Aosta, Sardegna, Campania, Bolzano; the fourth community is formed by Marche, Toscana, Piemonte; the fifth community is formed by Sicilia, Liguria,Puglia, Abruzzo, Umbria, Trento,, Calabria, Lazio, Friuli.

In the observation period, the number of extracted communities further decreases, see Figure 18(c) and there are: (i) Piemonte, Marche, Toscana; (ii) Lombardia, Veneto, Emilia, (iii) Basilicata, Molise, (iv) Puglia, Friuli, Valle d’ Aosta, Lazio, Abruzzo, Umbria, Campania, Trento, Liguria Bolzano, Calabria, Sardegna, Sicilia.

Figure 19 reports the evolution of Discharged/Healed Network Communities. It is possible to notice that in the first week, Figure 19(a), there are: a large community formed by Umbria, Piemonte, Toscana Campania, Sicilia, Sardegna, Abruzzo, Umbria, Calabria, Basilicata, Bolzano, Emilia, Valle d’Aosta, Friuli, Trento, Molise; a small community formed by Lombardia, Marche, Lazio; and single communities formed by Veneto. After three weeks the number of extracted communities increases, see Figure 19(b). In fact, Lombardia leaves the previous community and becomes a single one; Lazio, Emilia, Liguria and Veneto get together to form a second community; the third is composed by Friuli, Sicilia, Toscana; the last one is formed by Sardegna, Valle d’Aosta, Marche and Piemonte, Umbria, Campania, Molise, Abruzzo, Trento, Puglia, Calabria, Bolzano, Basilicata.

**Figure 19:**
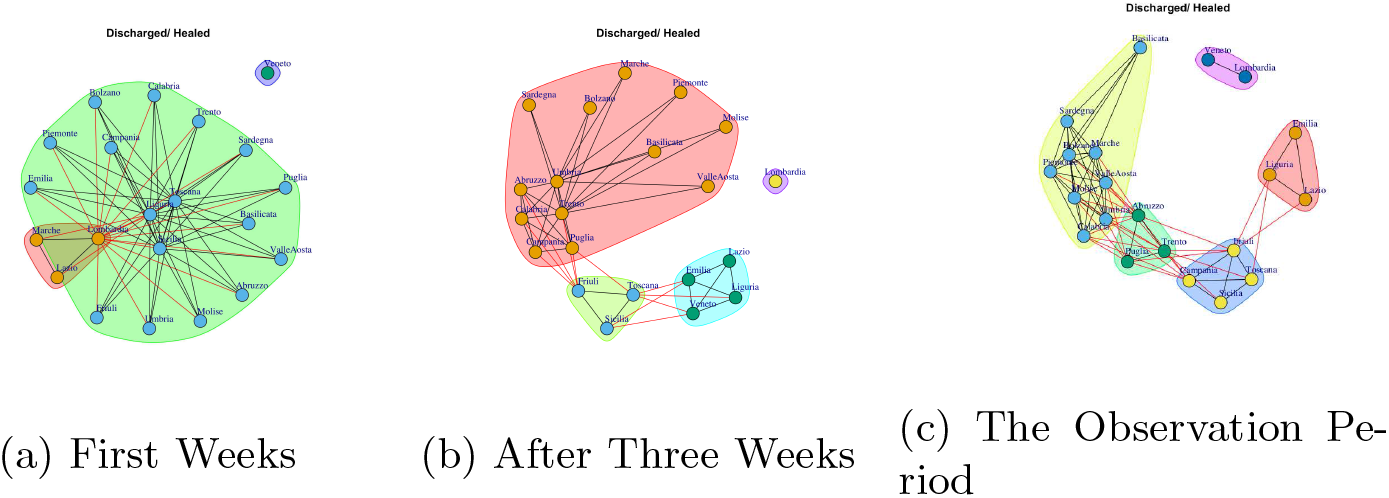
Evolution of Discharged/ Healed Network Communities.

Finally, Figure 19(c) shows the communities in the observation period. It is possible to notice five community. The first one consists of Lombardia and Veneto; the second one is composed by Emilia, Liguria, Lazio; the third one is composed by Friuli, Campania, Toscana, Sicilia; the fourth one is formed by, Puglia, Abruzzo, Trento; the last one is composed by Umbria, Sardegna, Calabria, Piemonte, Marche, Valle d’Aosta, Bolzano, Basilicata, Molise.

Figure 20 reports the evolution of Deceased Communities. Figure 20(a) reports the mined communities at the end of first week. It is possible to notice that there are: a large community formed by Emilia, Piemonte, Liguria, Campania, Abruzzo, Puglia, Valle d’Aosta, Umbria, Calabria, Sicilia, Campania, Trento, Lazio, Sardegna, Bolzano, Basilicata and Molise, Friuli; two single communities represented by Lombardia and Veneto; and a single community composed by Marche and Toscana.

**Figure 20:**
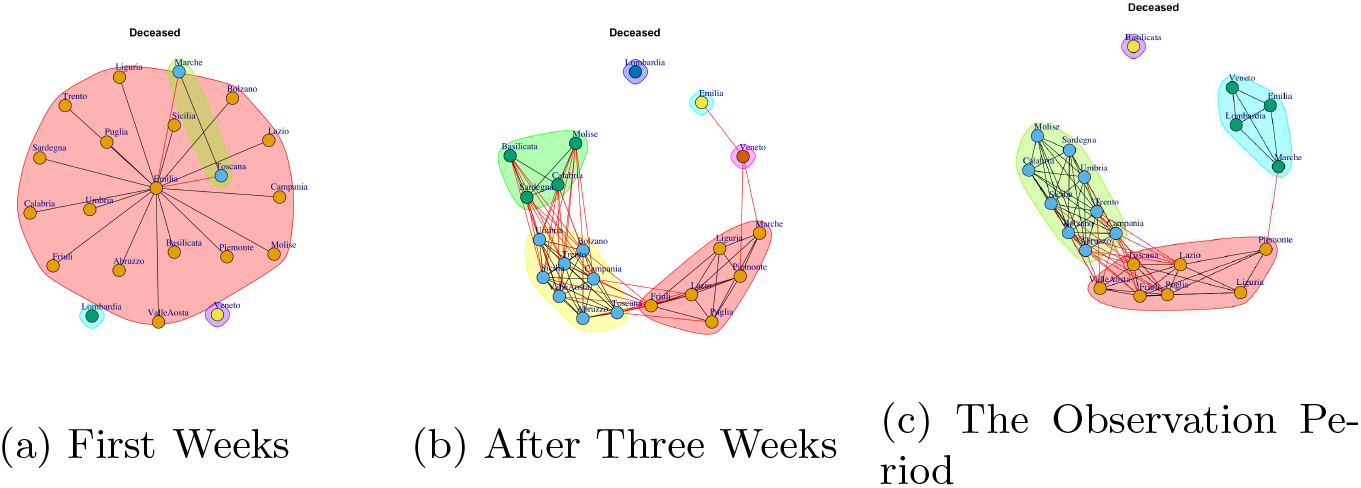
Evolution of Deceased Network Communities.

After three weeks, the number of extracted communities increase. In fact, the regions that form them vary by forming new communities, see Figure 20(b). The first community is composed by Lombardia that remains a single community; the second community is composed by Veneto and the third one is composed by Emilia; the fourth community is formed by Basilicata, Molise, Sardegna, Calabria; the fifth community is formed by Sicilia, Liguria,Puglia, Abruzzo, Umbria, Trento, Lazio, Friuli, Valle d’Aosta, Campania, Bolzano; Marche, Toscana, Piemonte.

In the observation period, the number of extracted communities decreases, see Figure 20(c) and there are: (i) Basilicata represents a single community, (ii) Piemonte, Toscana Liguria, Lazio, Friuli, Puglia, Valle d’ Aosta, (iii) Lombardia, Veneto, Emilia, Marche, (iv) Sicilia, Molise, Abruzzo, Umbria, Campania, Trento, Bolzano, Calabria, Sardegna.

Figure 21 reports the evolution of Total Cases Network Communities.

**Figure 21:**
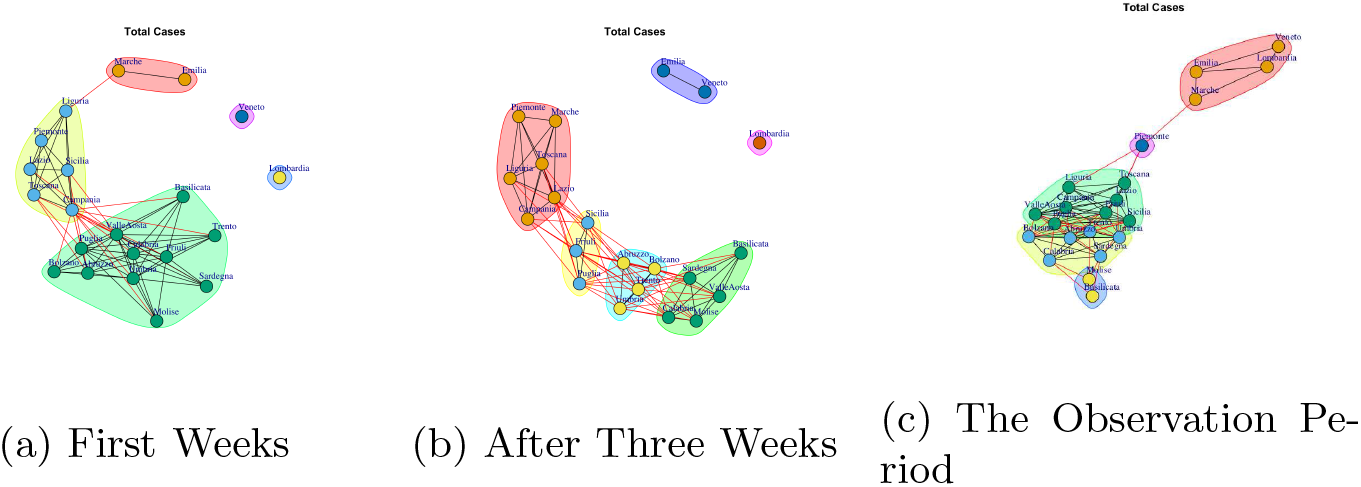
Evolution of Total Cases Network Communities

Figure 21(a) shows the five mined communities. The fist community is composed by Liguria, Toscana Lazio, Piemonte, Campania, Sicilia; the second one is formed Sardegna, Abruzzo, Umbria, Calabria, Basilicata, Bolzano, Valle d’Aosta, Friuli, Trento, Molise, Puglia; the third module comprises Marche, Emilia; the forth one Lombardia and the last one Veneto.

After three weeks (see Figure 21(b)) the communities are six. Veneto becomes a community among with Emilia; Marche moves in the community with Liguria, Toscana Lazio, Piemonte, Campania; and three new modules are formed: the first one is composed by Sicilia, Friuli and Puglia, the second one is composed by Abruzzo, Bolzano, Trento and Umbria, the third one is formed by Basilicata, Sardegna, Valle d’Aosta, Calabria, Molise.

At the end of the observation period the communities extracted are five. Figure 21(c) reports the communities. The first community is composed by Lombardia, Veneto, Emilia, Marche; the second community is composed by Piemonte; the third community is composed by Basilicata, Molise; the fourth community is composed by Toscana Liguria, Lazio, Campania, Friuli, Sicilia Puglia, Valle d’ Aosta formed, whereas Abruzzo, Umbria, Trento, Bolzano, Calabria, Sardegna formed the fifth community.

Figure 21 reports the evolution of Swab Network Communities. At the end of first week the mined communities are eight and they are reported in Figure 22(a). The fist community is composed by Umbria, Calabria, Valle d’Aosta, Trento, Sicilia, Abruzzo, Puglia; the second one is formed by Sardegna, Basilicata, Molise, Bolzano; the third module comprises Friuli, Campania, Liguria; Piemonte represents the forth community; the fifth one consists of Toscana and Lazio; the sixth community is represented by Emilia and Marche; the seventh is represented by Lombardia and the last one consists of Veneto.

**Figure 22:**
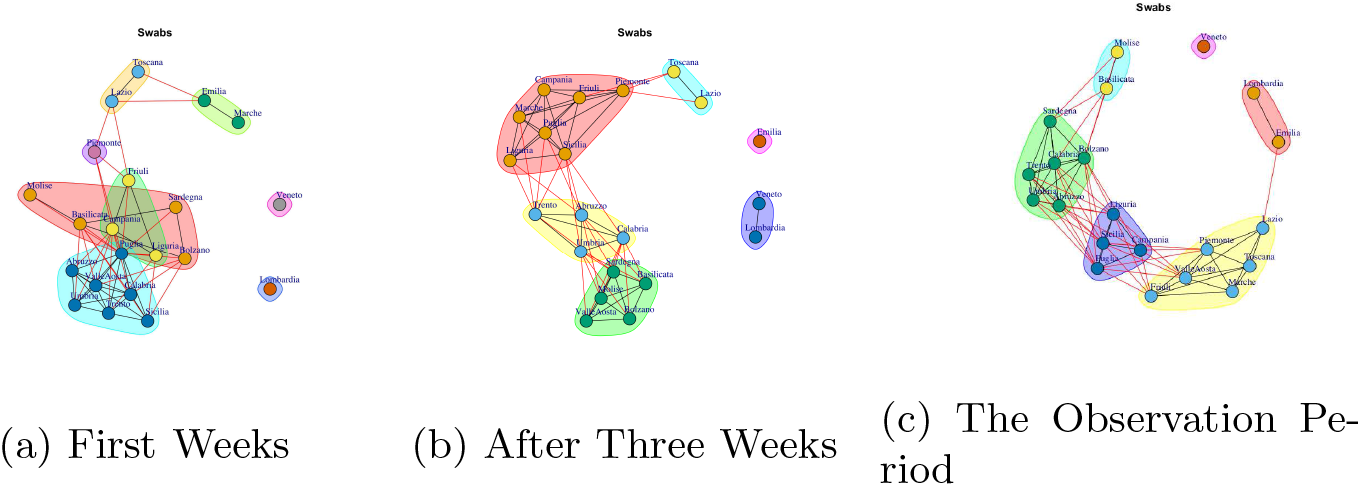
Evolution of Swabs Network Communities.

After three weeks, the number of communities (see Figure 22(b)) decreases. In fact, it is possible to notice six subgraphs. Emilia leaves the previous community and forms a single one. Lombardia and Veneto join together. Toscana and Lazio continue to form a community; Piemonte, Friuli, Campania, Marche, Puglia, Liguria, Sicilia form a forth community; Trento, Abruzzo, Umbria, Calabria, form a fifth community and the last one is composed by Sardegna, Basilicata, Molise, Bolzano, Campania, Valle d’Aosta;

In the observation period, the extracted communities remain six but the regions that form them vary, see Figure 22(c) The first community is composed by Veneto; the second community is composed by Lombardia, Emilia; the third community is composed by Basilicata, Molise; the fourth community is formed by Marche, Toscana, Lazio, Piemonte, Friuli, Valle d’ Aosta; the fifth community is formed by Sicilia, Campania, Liguria Puglia; while Abruzzo, Umbria, Trento, Bolzano, Calabria, Sardegna formed the sixth community.

In conclusion we can affirm that the topology of the communities varies, i.e. the regions join and leave them along time and the community consistency changes along time and with respect to the different available data.

## 4 Conclusion

The COVID-19 disease has spread worldwide in a matter of weeks. In Italy, the epidemic of COVID-19 has started in the north of the State and it quickly involved all regions. In this paper we evaluated the evolution of Italian COVID-19 data daily provided by the Italian Civil Protection. The goal this work is the network-based representation of COVID-19 diffusion similarity among regions and graph-based visualization with the aim to underline similar diffusion regions. We identified similar Italian region with respect to the available COVID-19 data and we mapped these ones in different networks. Finally, we performed a network-based analysis to discover communities of regions that show similar behaviour. As future work we plan to extend the study by adding other data such as particulate matter (PM10) data available at regional level.

## Data Availability

Raw data are available at: https://github.com/pcm-dpc/COVID-19
Similarity tables are provided in Supplementary Material.

## Acknowledgements

We thank Italian Civil Protection Department for freely providing online COVID-19 data.

